# A Metabolic Biomarker Panel for Congenital Heart Disease Assessment with Newborn Dried Blood Spots

**DOI:** 10.1101/2023.08.01.23293520

**Authors:** Scott R. Ceresnac, Yaqi Zhang, Kuo Jung Su, Qiming Tang, Bo Jin, James Schilling, C. James Chou, Zhi Han, Brendan J. Floyd, John C. Whitin, Karl G Sylvester, Henry Chubb, Ruben Y. Luo, Lu Tian, Harvey J. Cohen, Doff B. McElhinney, Xuefeng B. Ling

## Abstract

**Background:** Congenital heart disease (CHD) represents a significant contributor to both morbidity and mortality in neonates and children. The prompt recognition of CHD can facilitate timely and appropriate intervention, reducing the probability of complications and enhancing the prognosis for impacted newborns. However, unlike other rare conditions routinely identified through federal and state newborn screening (NBS) programs, there’s currently no analogous dried blood spot (DBS) screening for CHD immediately after birth.

**Objective:** This study was set to identify reliable metabolite biomarkers with clinical relevance, with the aim to assess feasibility of screening and subtype classification of CHD utilizing the DBS newborn screening method.

**Methods:** We assembled a cohort of DBS datasets from the California Department of Public Health (CDPH) Biobank, encompassing both normal controls and three pre-defined CHD categories (tetralogy of Fallot, inherited arrhythmia syndrome, neonatal cardiomyopathy). A robust, DBS-oriented metabolomic method, employing both global and targeted strategies based on liquid chromatography with tandem mass spectrometry (LC-MS/MS), was developed. To verify the reliability of this metabolic profiling, we conducted a correlation analysis comparing the absolute quantitated metabolite concentration in DBS against the CDPH NBS records. Additionally, for hydrophilic and hydrophobic metabolites, we executed significant pathway and metabolite analyses respectively. Finally, logistic and LightGBM models were established to aid in CHD discrimination and classification.

**Results:** Our metabolomic workflow demonstrated consistent and reliable quantification of metabolites in DBS samples stored at the California Department of Public Health (CDPH) for up to 15 years. Through this process, we discerned dysregulated metabolic pathways in CHD patients, including deviations in lipid and energy metabolism, as well as oxidative stress pathways. Furthermore, we identified three metabolites as potential biomarkers for CHD assessment, and an additional twelve metabolites as potential markers for classifying different CHD subtypes within DBS samples.

**Conclusions:** This study represents the first attempt to validate metabolite profiling results using long-term storage DBS samples procured from the high-quality conditions of the CDPH biobank. The results unveil distinct metabolic discrepancies between various CHD subtypes and healthy controls. Furthermore, our findings highlight the potential clinical applications of our DBS-based methods for CHD screening and subtype classification.

## Introduction

Congenital heart disease (CHD) ranks as the most prevalent form of congenital anomalies^1, 2^, making it a significant contributor to morbidity and mortality ^3, 4^, among neonates and children. With its birth prevalence on an upward trend, CHD has evolved into a major global health concern.

The importance of early diagnosis is paramount for the effective CHD intervention and treatment. Prenatal diagnosis and early detection advancements have contributed to a gradual decline in the mortality rate associated with CHD in children^5^. Various screening approaches for the detection of cyanotic congenital heart disease have been implemented and examined, including neonatal pulse oximetry screening^6^ and universal ECG screening^7, 8^. However, these methodologies exhibit less than 75% sensitivity in detecting critical CHD, presenting limitations in the detection of an array of congenital malformations, cardiomyopathic disorders, and inherited arrhythmia syndromes. These conditions contribute significantly to neonatal mortality and sudden death in children and adolescents^9^. Currently, there is no comprehensive, cost-effective screening method available at birth that can reliably and consistently detect the diverse range of CHD conditions that could potentially harm infants and children later in life.

Over the past decade, the application of cardiac-specific biomarkers, proteomic and metabolite analysis, mRNA, and small molecule screens has seen a significant increase^10–12^. The discovery of biomarkers associated with cardiac dysfunction, myocardial cell damage, and heart-specific tissue damage heralds a shift in our approach to screening, assessing, and treating cardiac diseases. It has been shown that structural abnormalities, such as the metabolism of acylcarnitines^13^, can create detectable metabolomic disturbances. Physiological alterations linked to cyanosis in cyanotic congenital heart disease, hemodynamic changes tied to volume and pressure loading in non-cyanotic lesions, tissue injury due to perfusion abnormalities in cardiomyopathic states, and cellular changes related to potassium, calcium, and sodium exchange defects in channelopathies, could all lead to distinguishable differences in the metabolomic profiles of neonates and children with cardiac disease^14–18^.

Routine neonatal screening serves as a tool to identify diseases that could potentially impact a child’s long-term health or survival. Each year, millions of infants in the United States undergo newborn screening (NBS), where substances in dried blood spots (DBS) are measured to check for certain genetic, endocrine, and metabolic disorders^19^. Early detection, diagnosis, and intervention can avert death or disability, enabling children to reach their full potential. Despite this, no DBS newborn screening exists for CHD at birth.

Our core hypothesis proposes that deep phenotyping of DBS metabolites at birth through liquid chromatography-mass spectrometry (LC-MS) could model and assess cardiac and other organ anomalies with high precision. This hypothesis was previously tested with our DBS analyses assessing other neonatal diseases^20^.

In this study, we developed an LC-MS based metabolic screening method to construct a baseline for neonate DBS metabolites and identify a DBS biomarker panel as a molecular surrogate to assess congenital cardiac abnormalities. Our findings attest to the robustness of our DBS-based LC-MS workflow, supporting the idea that assessing DBS metabolomic biomarkers may present a cost-effective and sturdy approach to evaluating the risk of CHD at birth. Further understanding the functional significance of these CHD biomarkers could provide fresh insights into the pathophysiology of CHD.

## Methods

### Ethics statement

This DBS NBS method development study involving human participants was reviewed and approved by ethics committees at Stanford University.

### Study design

The workflow for this study, as depicted in Fig. 1, was based on the collected dried blood spot (DBS) samples. Both global **hydrophilic/hydrophobic** and targeted liquid chromatography-tandem mass spectrometry (LC-MS/MS) metabolomic assays were conducted, adhering to the NBS DBS processing method. To verify the reliability of the metabolic profiling from samples that had been stored for years, a correlation analysis was carried out comparing the absolutely-quantified metabolite concentration with the California Department of Public Health (CDPH) gold standard. We identified significant metabolic pathways and features related to CHD and its subtypes. Finally, models for CHD diagnosis and subtyping were established to validate the effectiveness of the CHD-associated metabolic biomarker panel within the DBS samples.

**Figure 1.**
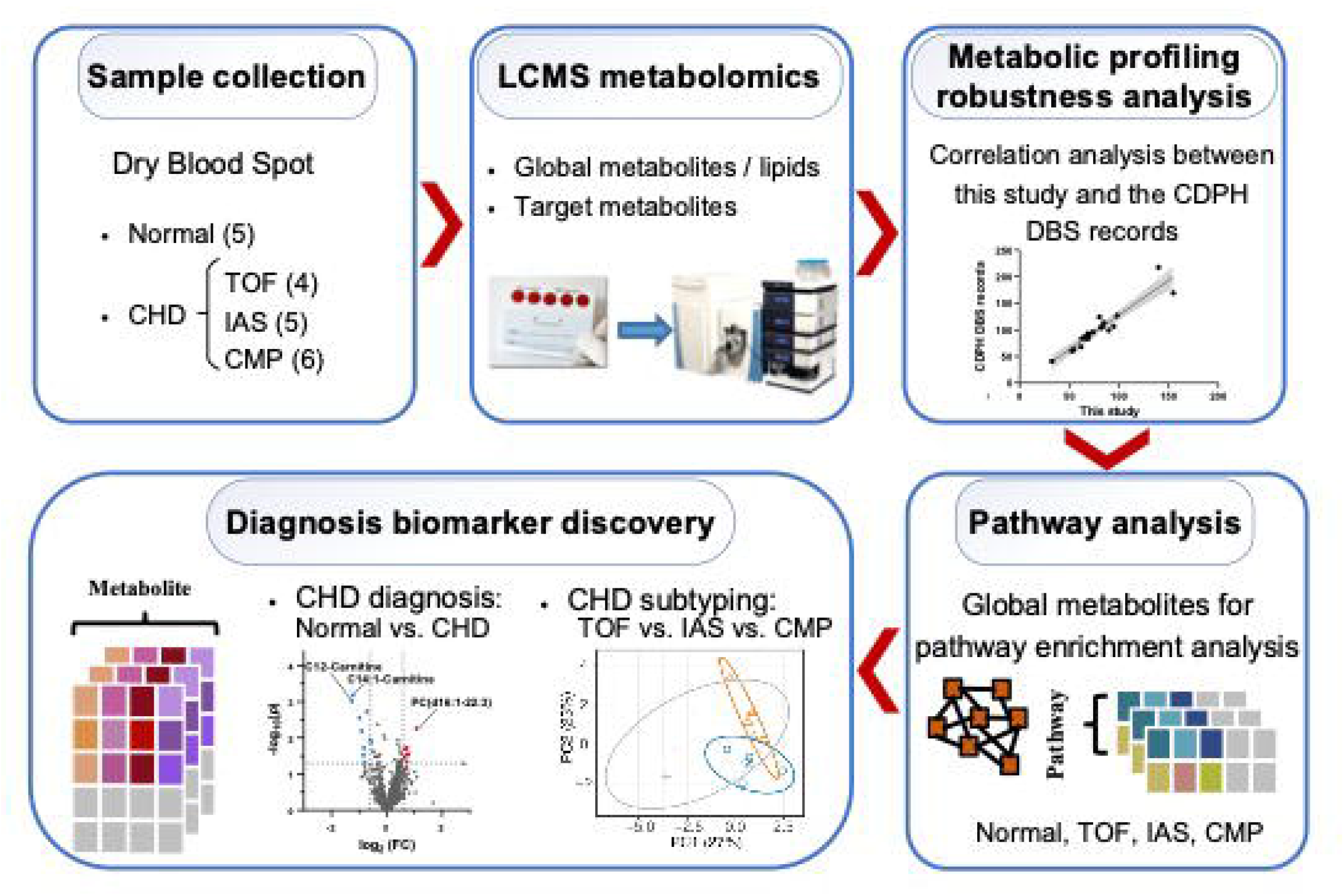
Study workflow diagram to apply metabolomic analytics to the neonate DBS samples and to discover CHD biomarkers. Abbreviations: CHD-Congenital heart disease, TOF-Tetralogy of Fallot, IAS-Inherited arrhythmias syndromes, CMP-Cardiomyopathies.

#### Cohorts

We randomly assembled a retrospective cohort of DBS profiling datasets from the CDPH Biobank. These DBS samples had been stored for up to 15 years since the time of newborn testing analysis, consented to be preserved at -20°C in the CDPH Biobank. In total, we included 20 clinical DBS samples, consisting of 5 healthy controls (HC) and 15 cases of CHD spanning different categories. The 15 CHD cases included 4 instances of Tetralogy of Fallot (TOF), 5 cases of inherited arrhythmia syndromes (IAS), and 6 instances of cardiomyopathies (CMP).

#### Sample preparation

The DBS samples were retrieved from -20°C freezers and thawed on ice. Each DBS sample was cut into 3mm diameter sections, and two of these sections were transferred into the same Eppendorf tube.

### Global hydrophilic and targeted metabolomics

We followed our previously developed standard operational protocols^21^ of global hydrophilic and targeted metabolomics for the workflow in this study, including sample preprocessing, mass spectrometry signal acquisition, quality control, data pre-processing, and metabolite biomarker structural identification.

For global hydrophilic metabolomics, a mixture of 250 μL extraction buffer (pre-chilled to -20 °C) consisting of methanol, acetonitrile and ddH2O (5:3:2 v/v) was added to each tube containing DBS punches. Samples were vortexed and centrifuged at 10,000 g for 10 min at 4°C. Supernatant (180 μL) from each sample was transferred into a clean Eppendorf tube. 100 µL of each sample extract was transfer into an auto-sampler vial for UHPLC-MS (Ultra-High Performance Liquid Chromatography) analysis.

For targeted metabolomics, a mixture of 200 µL methanol / acetonitrile (1:1, v/v) was added into the tube containing DBS punches. Samples were vortexed vigorously and centrifuged at 10,000 g for 10 min at 4°C. The supernatant of each sample was transferred into a new tube and dried under nitrogen stream. 10 µL of Internal Standard Solution, 90 µL of Extraction Buffer, and 200 µL hexane were added into the reconstituted tube for extraction. The sample was vortexed vigorously for 1 min and centrifuged at 12,000 g for 5 min. 180 µL of upper layer was transferred into another 1.5-mL centrifugal tube and dried under nitrogen stream again. The residue was reconstituted with 100 µL of Derivatization Buffer. The reconstituted sample was incubated at 95°C for 15 min. After derivatization, 100 µL of each reconstituted sample was transferred into an auto-sampler vial for the analysis of fatty acids. 80 µL of lower layer was transferred into an auto-sampler vial for the analysis of amino acids and acylcarnitines.

### Global hydrophobic metabolomics

For global hydrophobic metabolomics, a mixture of 400 μL of chloroform / methanol (1:1, v/v) and 200 µL of water with lithium chloride were added to each tube containing DBS punches. Afterwards, the sample was vortexed rigorously for 30 sec and centrifuged at 12,000 g for 5 min. The bottom layer of each sample was transfered into another tube. The top layer was re-extracted with 400 µL of chloroform and vortexed for 30 sec and centrifuged at 12,000 g for 5 min. The bottom layer was removed and combined with extract from previous trial. The combined extract was dried under nitrogen and reconstituted with 100 µL of methanol: chloroform (1:1, v/v). Thereafter, the DBS extract was transferred into auto-sampler vial with micro-insert for LC/MS analysis.

For global hydrophobic metabolomics, 5 μL of DBS extract was injected via a Vanquish UHPLC system. The mobile phase was methanol with 10 mM ammonium acetate at a flow rate of 0.1 mL/min for a total run time of 3 minutes. The conditions of ionization source were set at 3.4 kV for spray voltage, 15 for sheath gas, 5 for aux gas, 325°C for capillary temperature, 55 for S-lens, and 250°C for vaporizer temperature. The MS spectra were acquired with 2 scans using an AGC target of 1e6 and a resolution of 120,000 (FWHM at 200 m/z) from 200 to 1200 m/z. The column oven was maintained at 25°C throughout the analysis.

#### Metabolic pathway enrichment analysis with global hydrophilic and hydrophobic metabolomics

To carry out pathway enrichment analysis on both hydrophilic and hydrophobic mass spectrometric profiling results, a univariate analysis was utilized to compute the fold change and p-value (using Student’s t-test) for each component across the three different CHD subtypes. Following the correction for false discovery, components that showed significant changes (adjusted p-value < 0.05) were chosen for subsequent analysis.

All significantly altered hydrophilic metabolites were categorized into KEGG pathways^22^ for further examination, while significantly changed hydrophobic metabolites were grouped into the Lipid Map database^23^. Enriched pathways of significance were identified with a Fisher’s exact test p-value of < 0.05.

#### Statistic learning for multi-class classification

Orthogonal partial least squares discriminant analysis (OPLS-DA) was performed using global hydrophilic and hydrophobic metabolic profiling results. Unsupervising clustering results were used to visualize the two-dimensional clustering patterns of HC, TOF, IAS and CMP DBS samples. With OPLS-DA results, the performance of the discrimination between two of these DBS sample categories was assessed using a receiver operating characteristic curve (ROC) and the area under the curve (AUC).

To distinguish between CHD patients and healthy controls, a logistic model^24^ was constructed based on targeted metabolomic profiling results. For CHD subtyping, the importance of identified targeted metabolites was determined using a gradient boosting machine provided by the LightGBM library^25^. Analysis was implemented with default parameters, and the metric used for early stopping was set to the error rate for multi-class classification^26^. Metabolites were then ranked based on their normalized importance scores. Metabolites with cumulative importance greater than 80% were selected as biomarkers.

The significantly relevant biomarker metabolites associated with CHD diagnosis and subtyping were then proposed and discussed.

## Results

### Demographics and Clinical Characteristics

**Table 1** summarizes the cohort demographics and clinical characteristics. This includes a cohort of 20 neonates, comprising of 5 controls and 15 CHD patients. Of these 20 neonates, 11 were boys (55%) and 9 were girls (45%). The 15 CHD patients comprised 4 diagnosed with CHD-Tetralogy of Fallot (TOF), 5 with CHD-inherited arrhythmia syndromes (IAS (2 Brugada, 3 Long QT syndrome)), and 6 with CHD-cardiomyopathies (CMP (3 dilated, 3 hypergtrophic cardiomyopathy)). No statistically significant differences were observed in the distribution of gender and race across all groups.

**Table 1.**
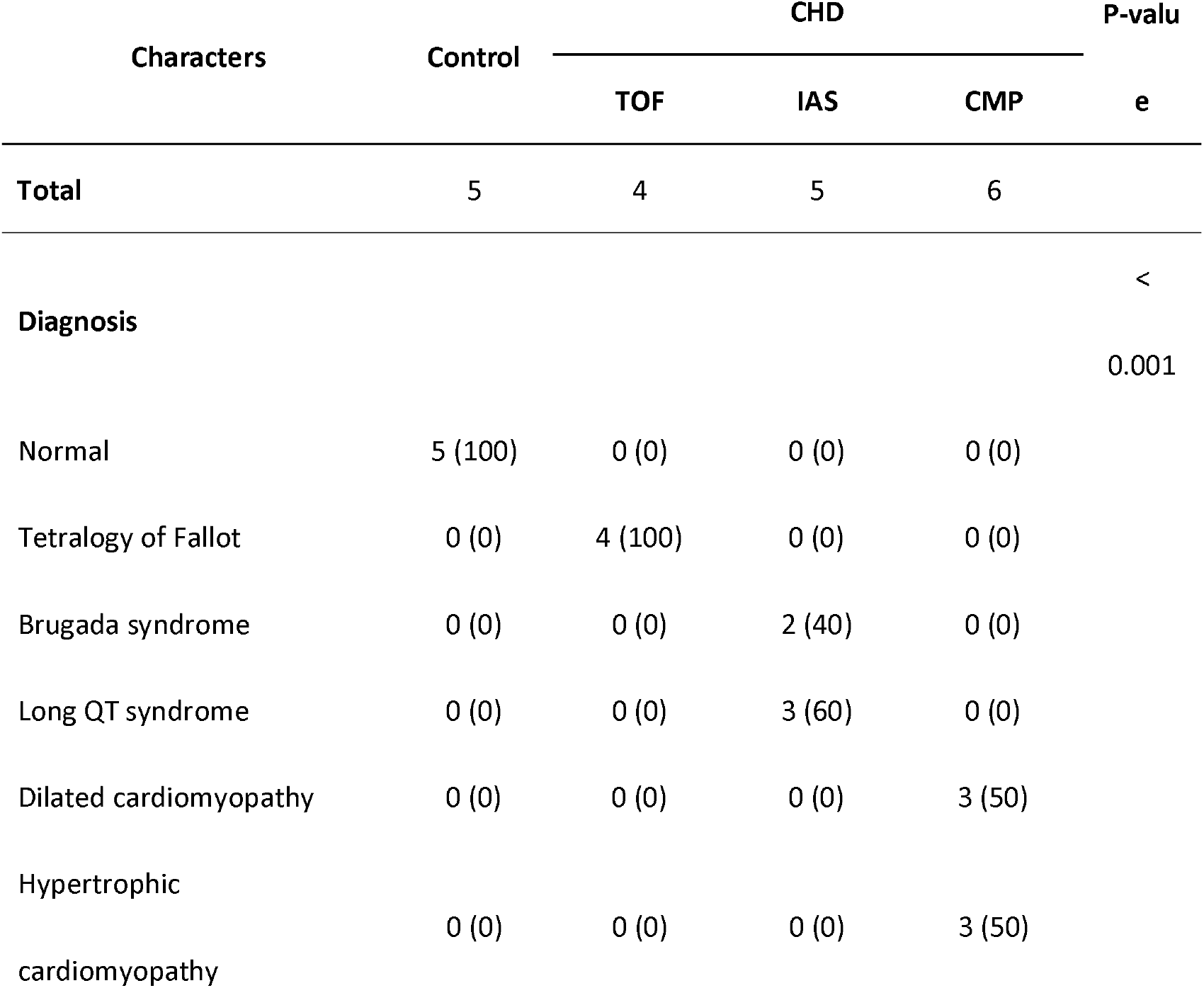
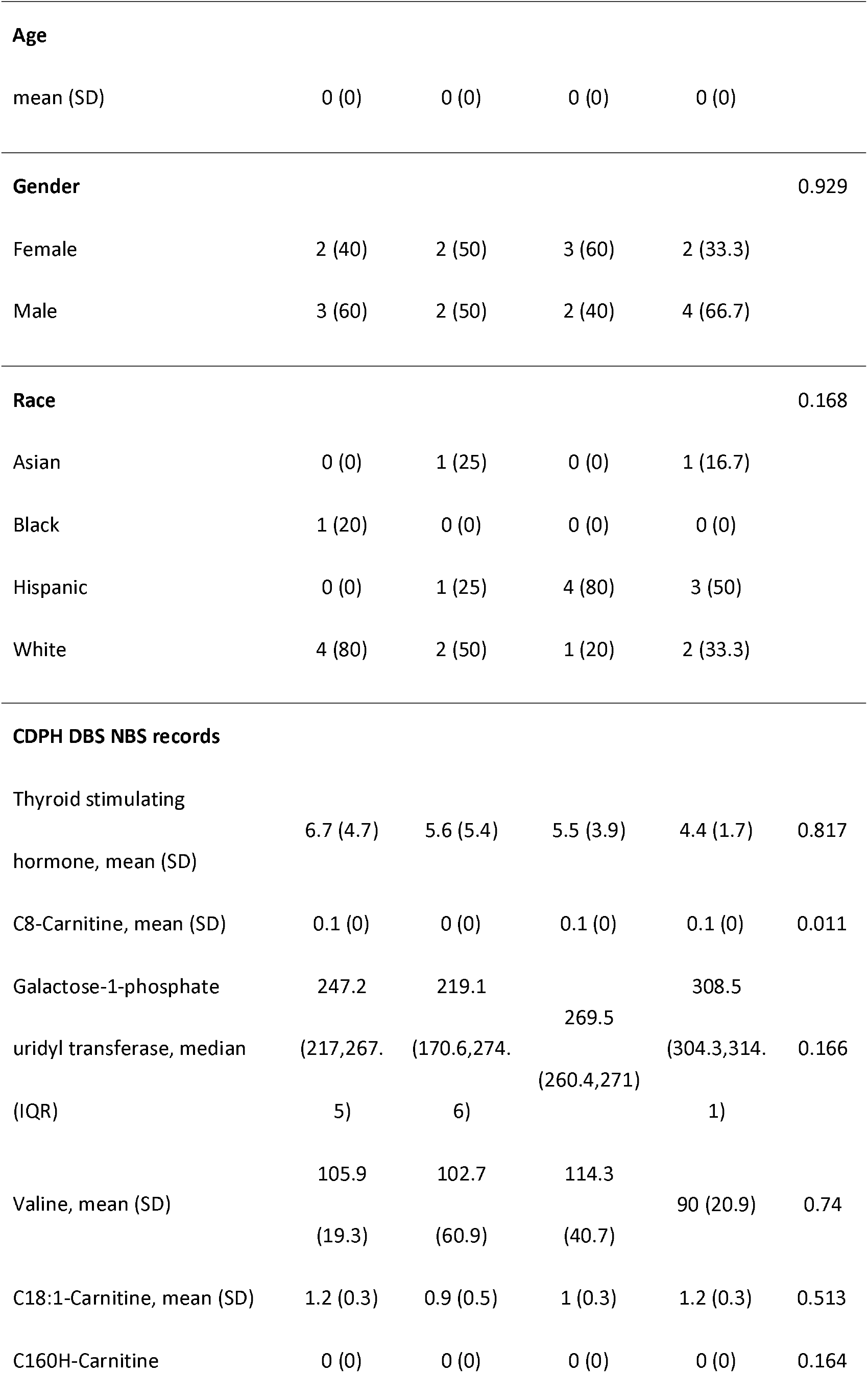
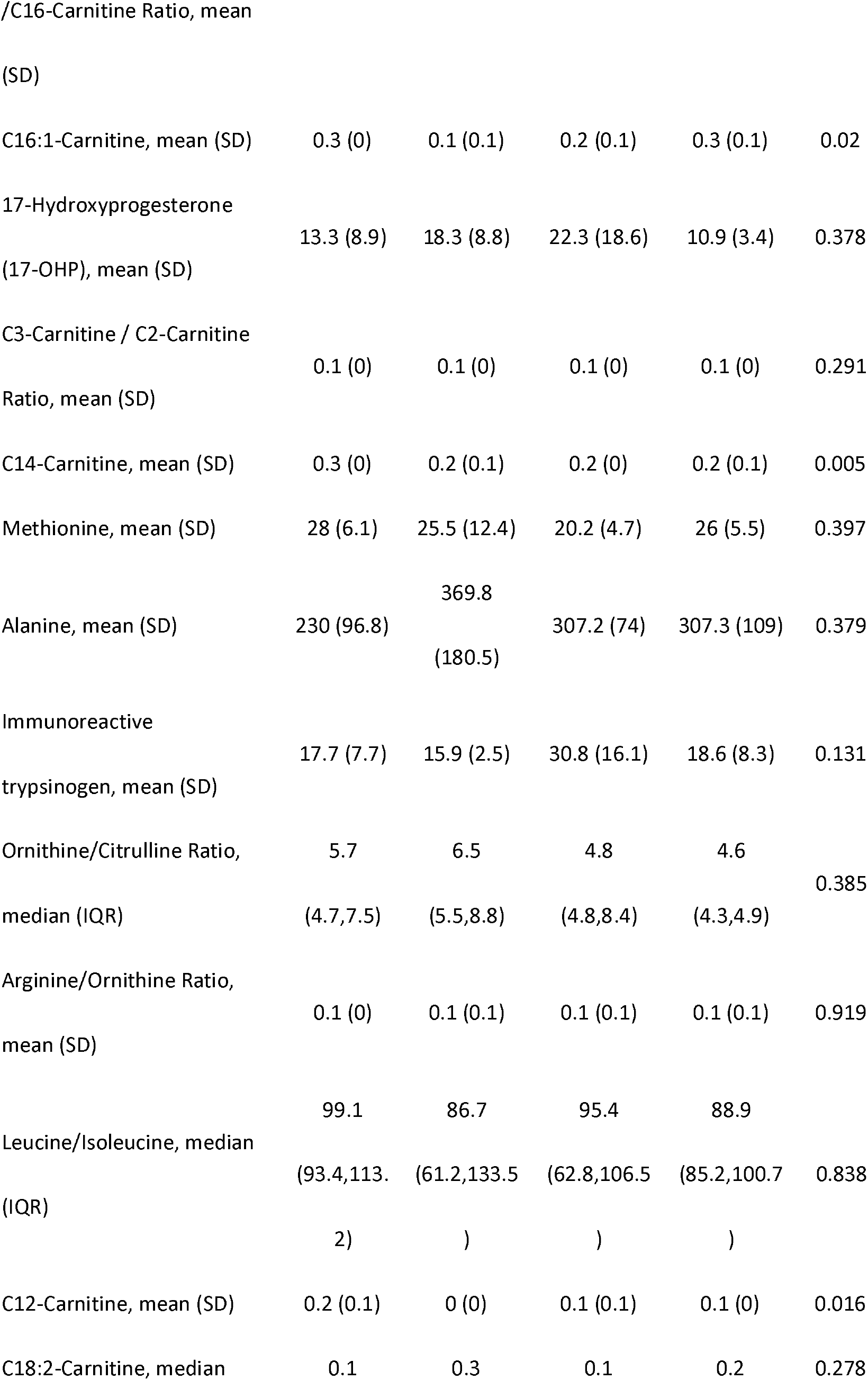
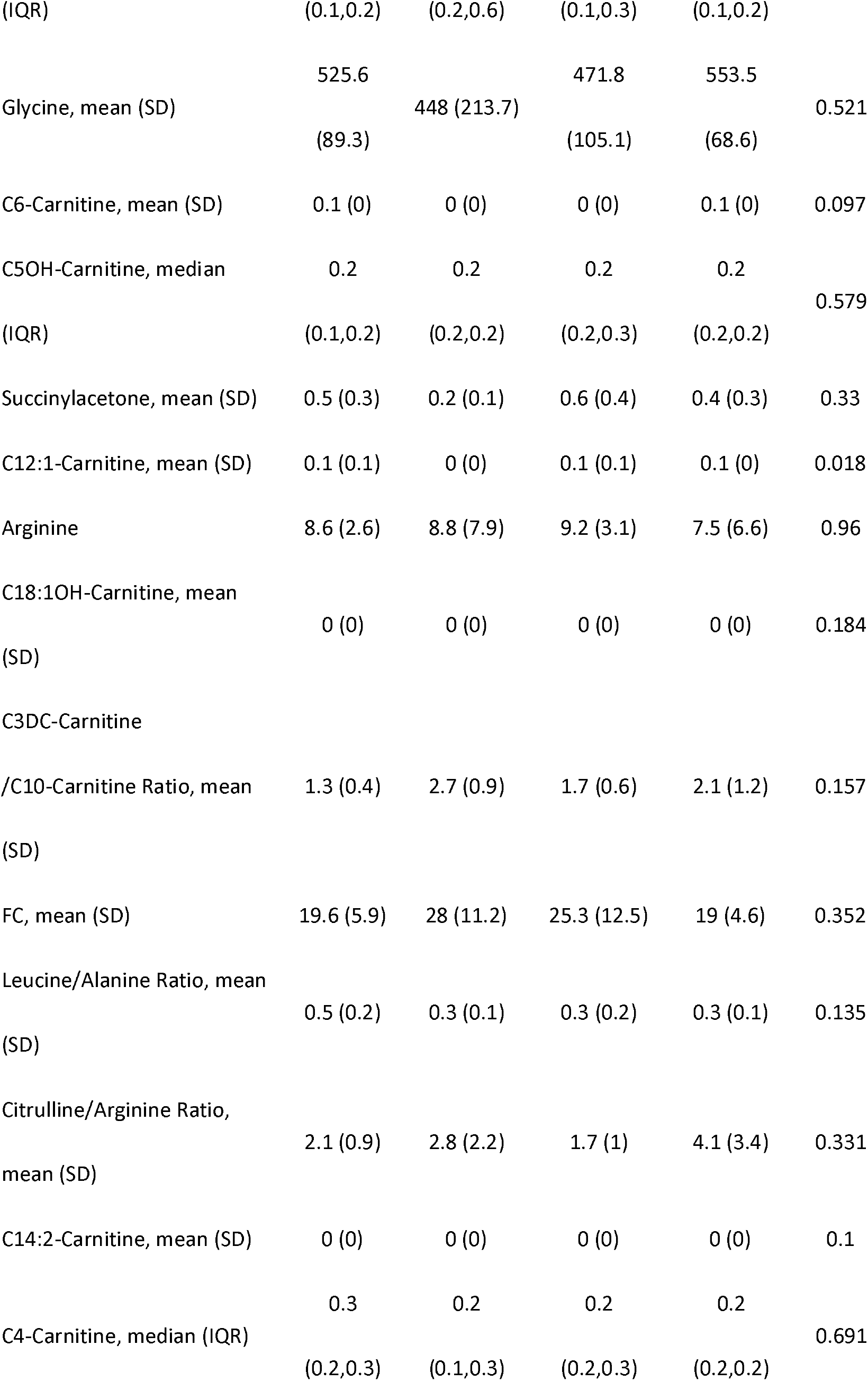
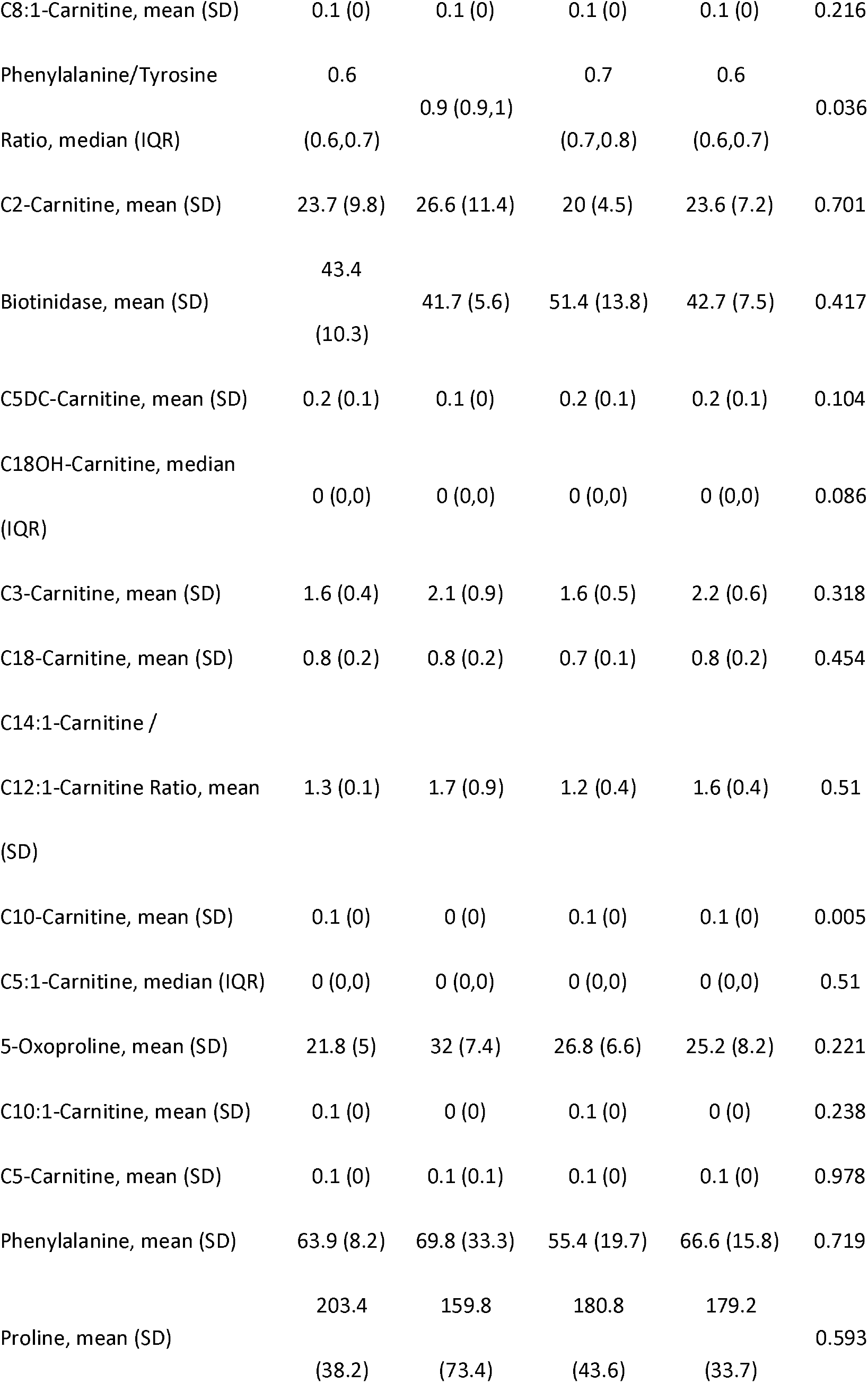
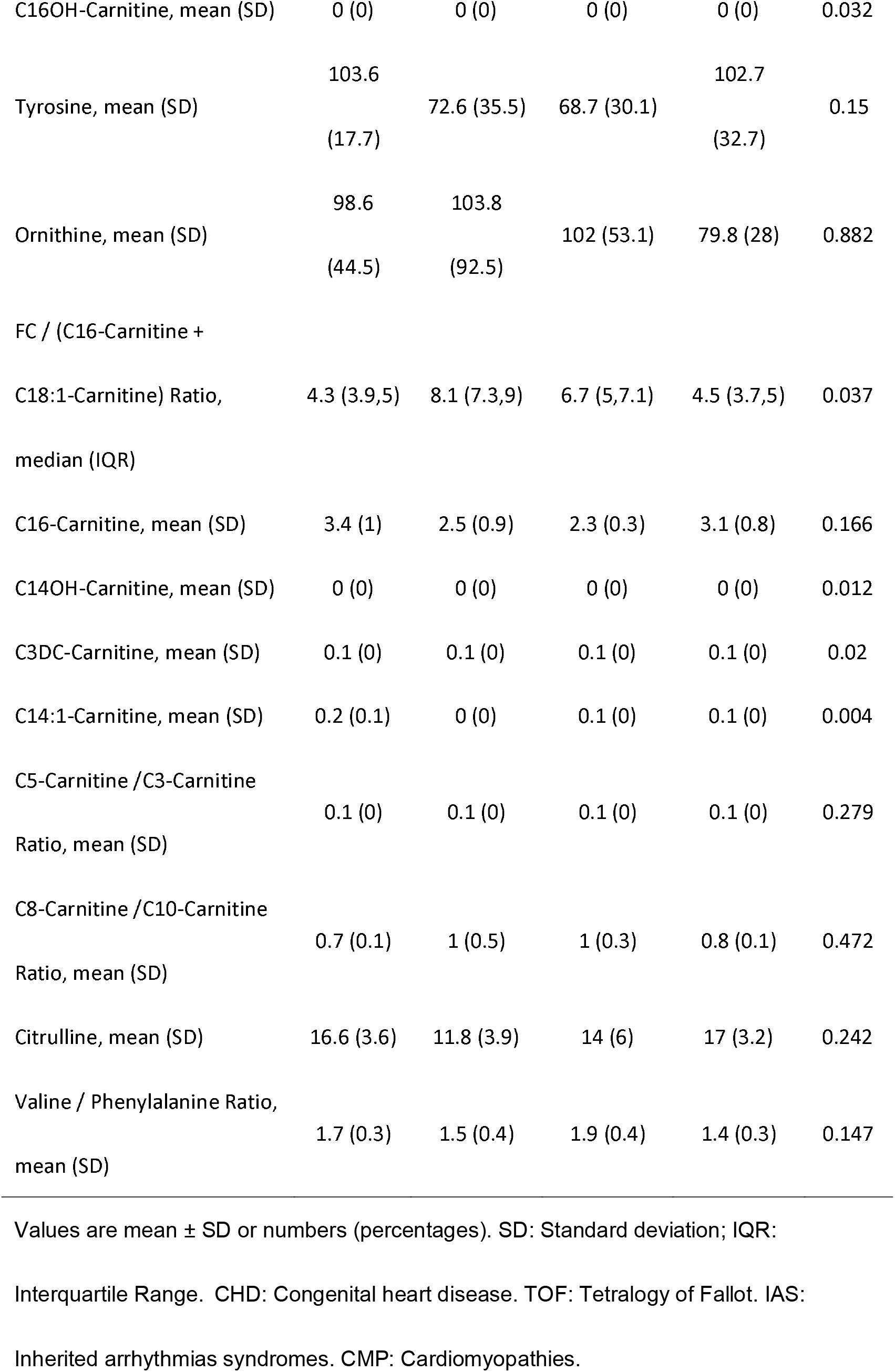
The demographics of CDPH DBS Samples.

The DBS samples had been stored in the CDPH biobank at -20 °C for up to 15 years, as shown in Fig. 2. We reanalyzed the concentrations of 28 metabolites, which are common to the CDPH DBS NBS records (Table 1) and include amino acids, free carnitines, and acylcarnitines. Fig. 3 shows that 24 out of the 28 metabolites exhibit a strong correlation, affirming both the robustness of our metabolomic profiling workflow and the reliability of these DBS samples even after many years of storage.

**Figure 2.**
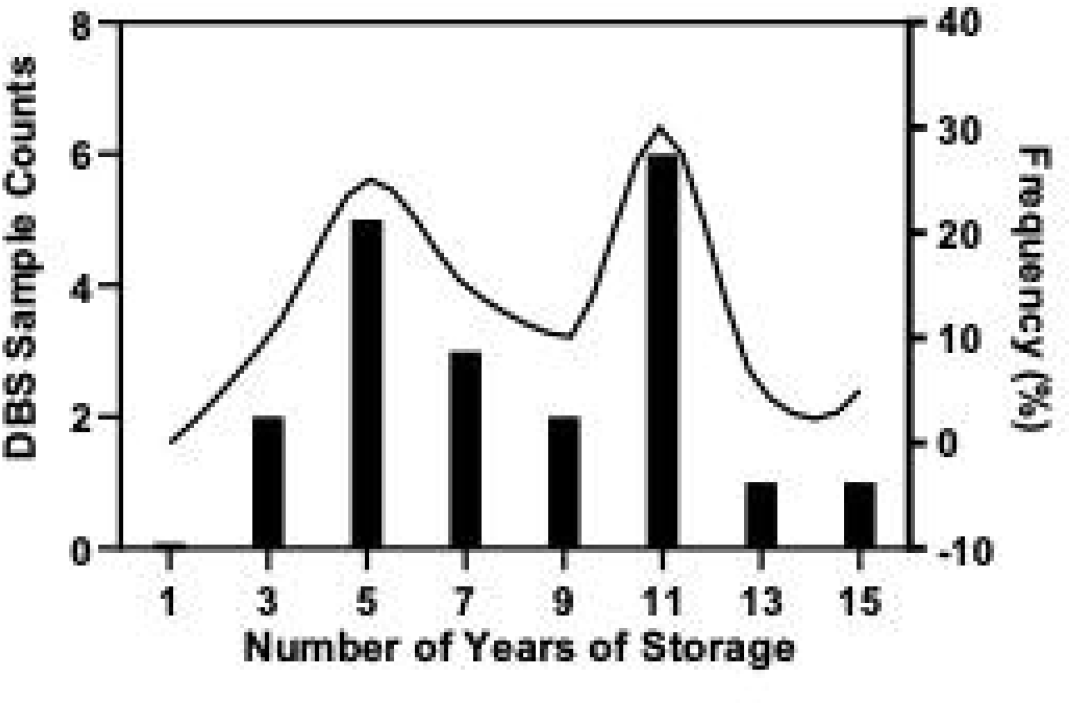
Statistical distribution of DBS Samples storage times in California Department of Public Health (CDPH) Lab.

**Figure 3.**
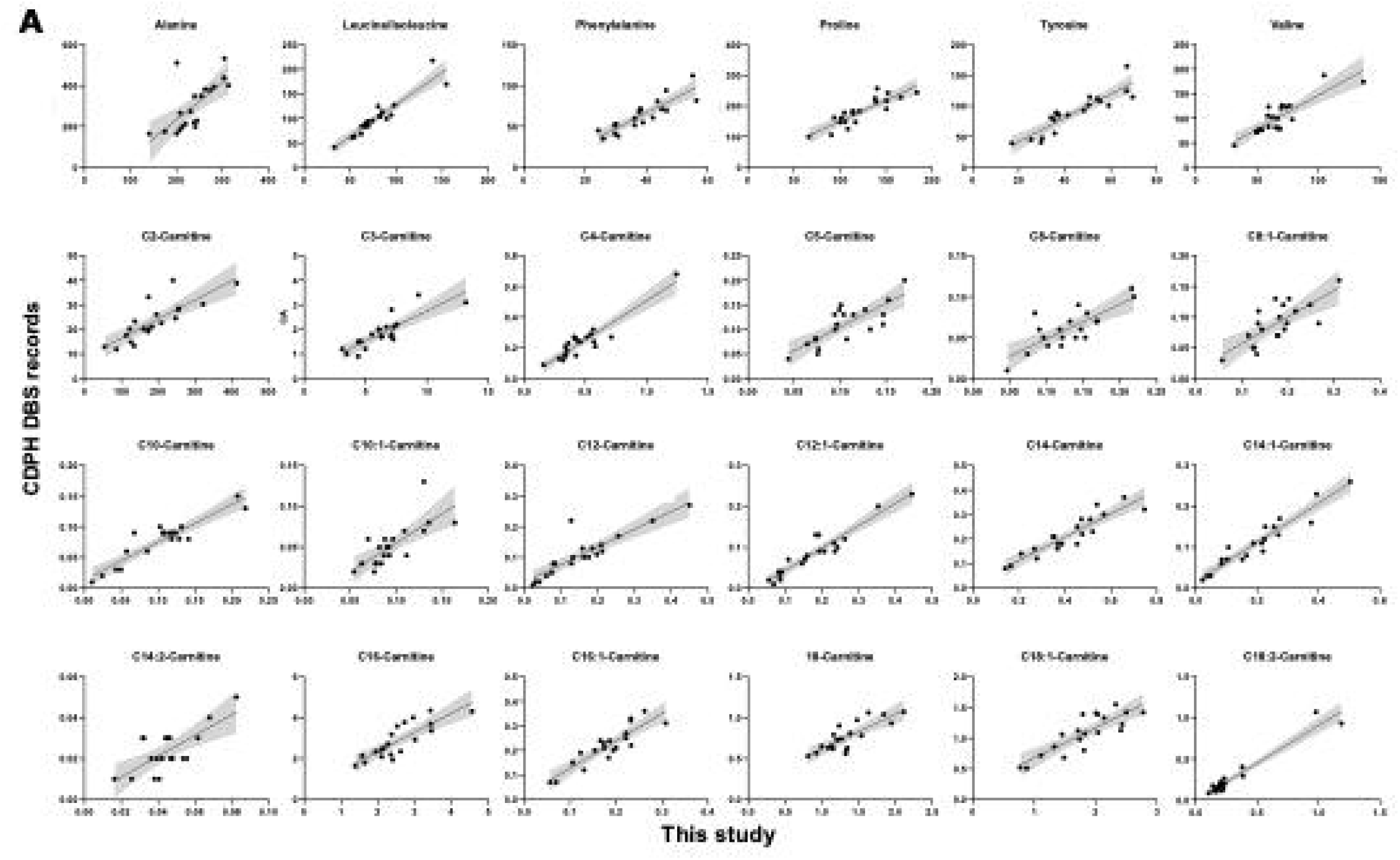
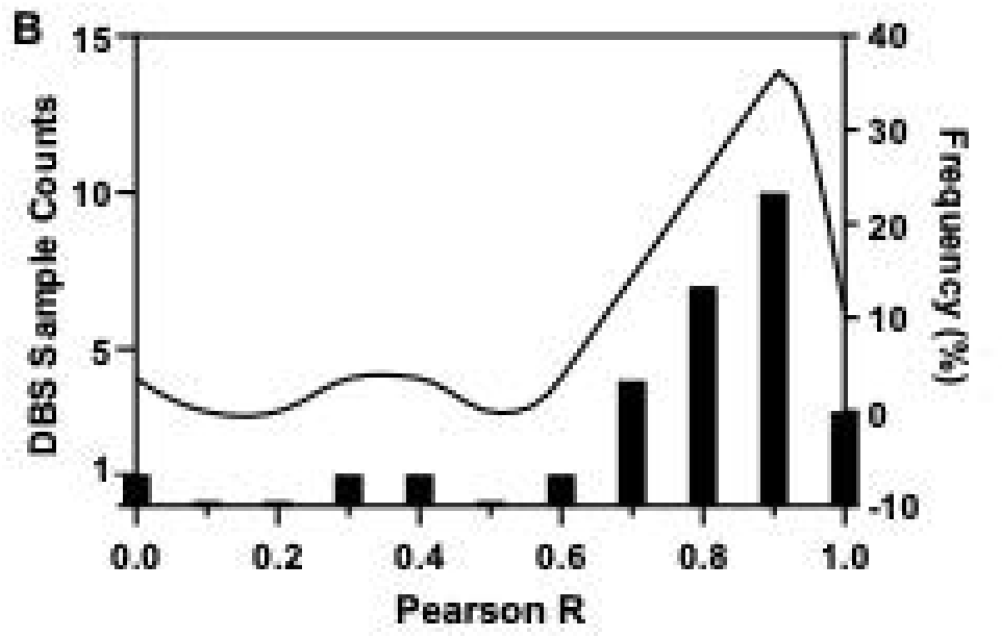
Impact of the time storage of metabolites. A) scatter plots showing the positive correlations of metabolic profiling between this study (X-axis) and the CDPH DBS records (Y-axis). B) Statistical distribution of the correlation coefficient.

### Exploration of the unique metabolic patterns among different CHD subtypes

Two distinct high-throughput global mass spectrometric workflows were conducted, focusing on hydrophilic and hydrophobic metabolites separately. After data preprocessing, a total of 1829 compounds were identified in the global hydrophilic metabolomic profiling, and 1383 compounds were identified in the global hydrophobic metabolomics.

Multivariate analysis, specifically sparse partial least squares discriminant analysis (OPLS-DA), was then performed on the global hydrophilic and hydrophobic metabolomics to investigate different CHD subtypes, including TOF, IAS, and CMP. The results, presented in **Fig. 4** and **Table 2**, revealed distinct clustering patterns for samples from various CHD subtypes in both the global hydrophilic and hydrophobic metabolic profiling.

**Figure 4.**
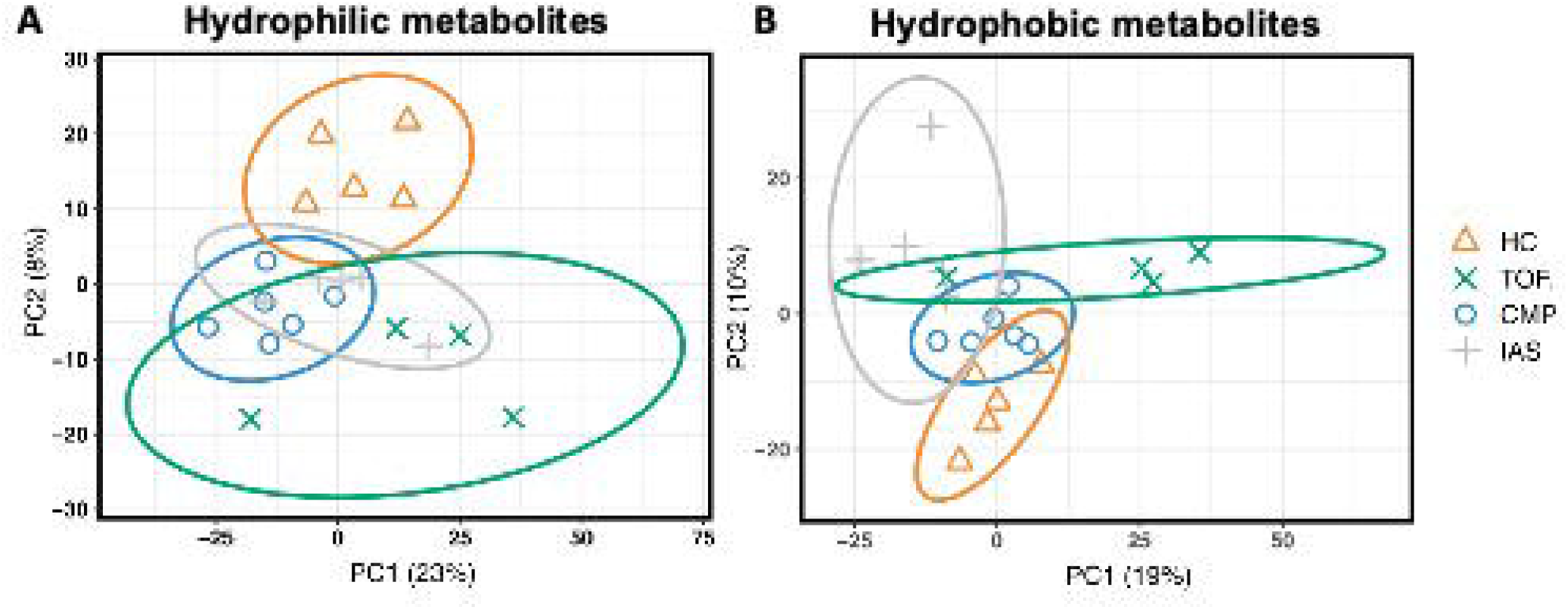
Orthogonal partial least squares discriminant analysis (OPLS-DA) using the global hydrophilic and hydrophobic metabolic profiling results of health control (HC), CHD-Tetralogy of Fallot (TOF), CHD-inherited arrhythmia syndromes (IAS) and CHD-cardiomyopathies (CMP). A) clustering results of hydrophilic metabolic profiling B) clustering results of hydrophobic metabolic profiling.

**Table 2.** Discernibility ability to CHD subtypes by global hydrophilic and hydrophobic metabolomics with orthogonal partial least squares discriminant analysis (OPLS-DA) .

|  | Hydrophilic |  | Hydrophobic |  |
| --- | --- | --- | --- | --- |
|  | AUC | P-value | AUC | P-value |
| HC vs Other(s) | 1.00 | 0.001 | 1.00 | 0.001 |
| CMP vs Other(s) | 0.881 | 0.008 | 0.607 | 0.458 |
| IAS vs Other(s) | 0.533 | 0.827 | 0.987 | 0.001 |
| TOF vs Other(s) | 0.969 | 0.005 | 0.906 | 0.014 |

HC: Health Control. TOF: Tetralogy of Fallot. IAS: Inherited arrhythmias syndromes. CMP: Cardiomyopathies.

Interestingly (**Fig. 4** and **Table 2**), CHD-IAS could not be reliably distinguished from other groups (ROC AUC, 0.607; P value, 0.458) based on the global hydrophilic metabolomics, while CHD-CMP also showed poor distinction from other groups (ROC AUC, 0.533; P value, 0.827) based on the global hydrophobic metabolomics. These findings suggest that each CHD subtype may indeed exhibit unique metabolic differences compared to both the healthy control and other CHD subtype groups.

#### Pathway analysis with global hydrophilic and hydrophobic metabolomics

To identify specific metabolic differences in CHD subtypes, pathway enrichment analysis was performed on the significant changes observed in global metabolism (adjusted P value < 0.05) (Fig. 5). For hydrophilic metabolites, the KEGG pathway database was utilized, while the Lipids Map database was used for hydrophobic metabolites. The results depicted in Fig. 5 show that the Arachidonic acid metabolism and Monoradylglycerols pathways are consistently and significantly enriched in all three CHD subtypes. Additionally, the Linoleic acid metabolism, serotonergic synapse, and sphingoid bases pathways are significantly enriched solely in the IAS and TOF subtypes of CHD. Moreover, Quinones and hydroquinones were found to be significantly enriched only in CHD-CMP, while Arginine and ornithine metabolism showed significant enrichment exclusively in CHD-IAS. These findings suggest the presence of diverse metabolic pathway changes among the different CHD subtypes, emphasizing the importance of considering both hydrophilic and hydrophobic metabolites when constructing a CHD diagnosis panel. Such a comprehensive approach will enable a more accurate and informative characterization of the metabolic profiles associated with different CHD subtypes.

**Figure 5.**
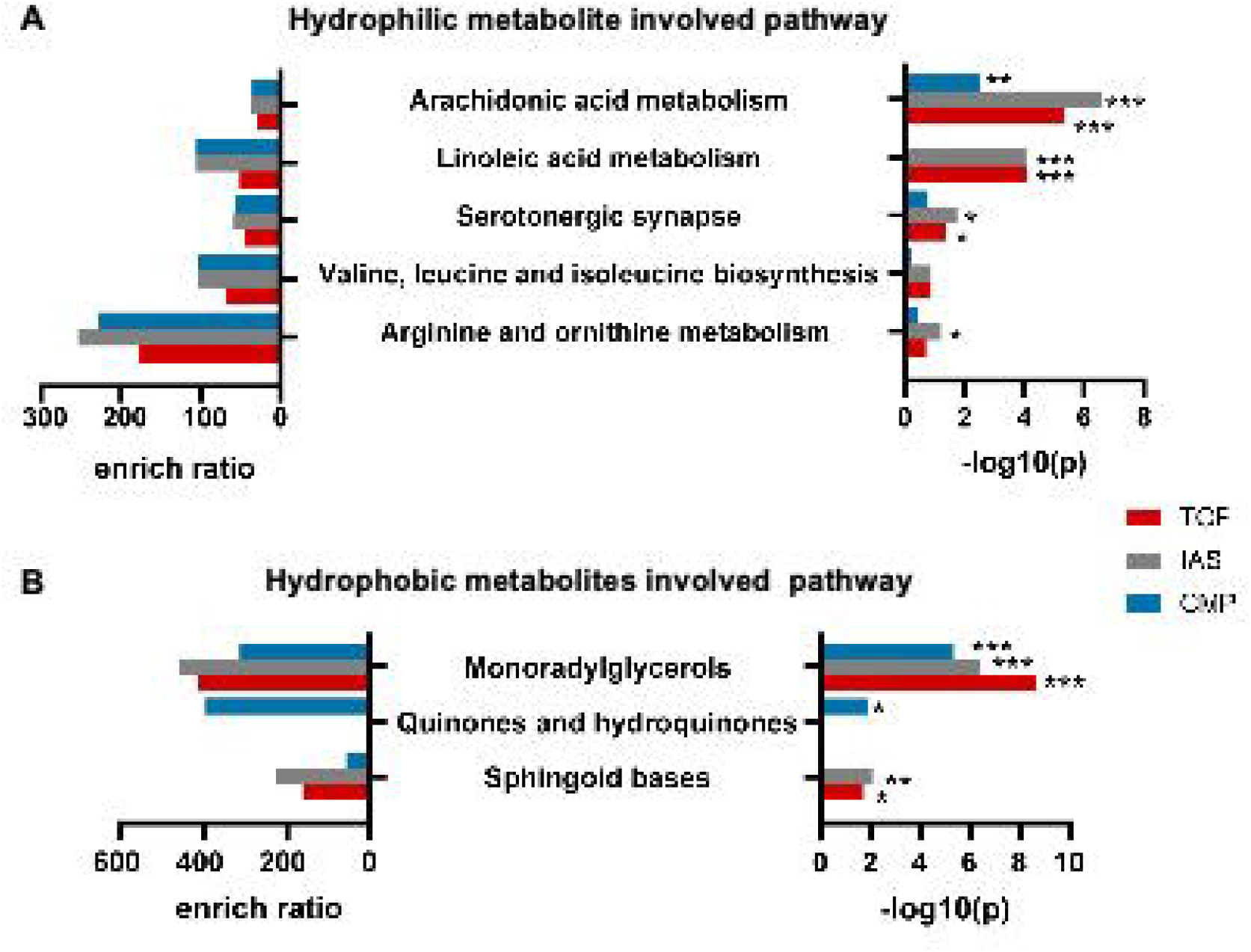
Significant Metabolic pathways altered in different CHD subtypes. Pathway enrichment analysis on the A) global hydrophilic and B) hydrophobic metabolic profiling. All significant changed components (P value <0.05, Student’s t-test) in CHD-Tetralogy of Fallot (TOF), CHD-inherited arrhythmia syndromes (IAS) and CHD-cardiomyopathies (CMP) are mapping to KEGG metabolic pathways and Lipid Map database, respectively. *: P value <0.05, **: P value <0.01, ***: P value <0.001.

### Identification of a metabolic signature for CHD

Using our targeted metabolomics methods, we conducted quantitative measurements of 836 targeted metabolites and lipids in DBS samples collected from both CHD patients and healthy controls. Upon comparing CHD patients with healthy controls, we identified 3 biomarker metabolites (p < 0.05) through univariate analysis, namely PC(d16:1-22:3), C14:1-Carnitine, and C12-Carnitine (**Fig. 6A** and **Fig. 6B**).

**Figure 6.**
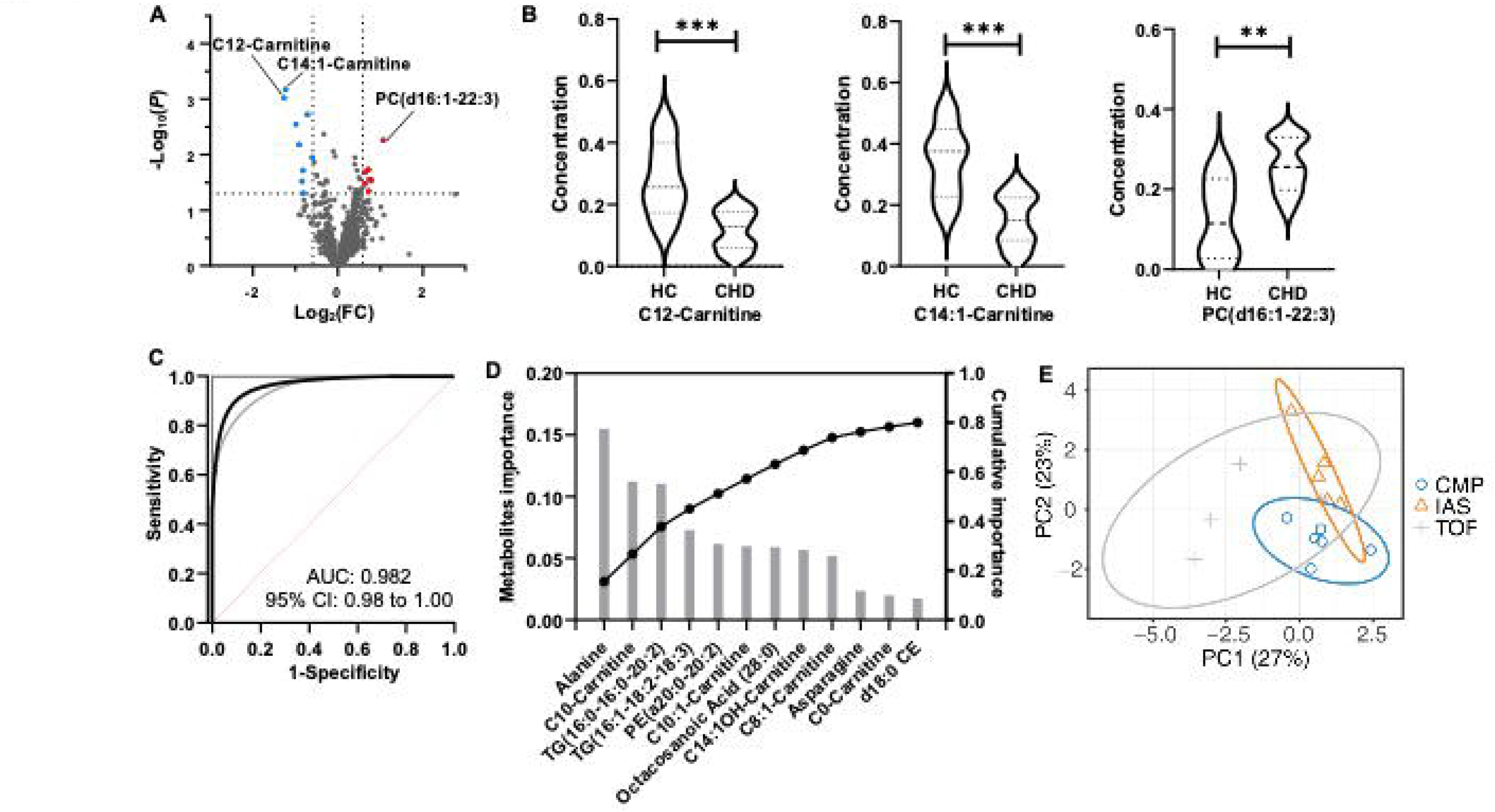
The application of targeted metabolomics to discover CHD diagnosis and subtyping biomarker metabolites using DBS samples. A) Volcano plots for screening significant changed metabolites associated with CHD, the metabolites with P value < 0.05 and fold change >1.5 are marked as red dots and the metabolites with P value < 0.05 and fold change < 0.67 are marked as blue dots. B) violin plots for three biomarker metabolites for CHD diagnosis. *: Student’s P value <0.05, **: P value <0.01, ***: P value <0.001. C) the smoothed receiver operating characteristic curve (AUC ROC) of logistic model based on three biomarkers. 95% confidence intervals are shown in grey lines. D) the importance score of the 12 metabolites associated with CHD subtyping which have an 80% cumulative importance in total. E) PLS-DA cluster results using 12 metabolites for CHD subtyping.

To further explore the diagnostic potential of these biomarkers, we constructed a logistic model based on their concentrations. This logistic model exhibited high accuracy in distinguishing between CHD patients and healthy controls, achieving an area under the curve (AUC) of 0.982 (95% CI: 0.92-1.00) (**Fig. 6C**). The sensitivity and specificity of this logistic model were 92.8% and 75.0%, respectively.

Regarding the metabolic signature for CHD subtyping, we employed the LightGBM model to calculate the importance of each metabolite in the multiclass classification of the three types of CHD. This analysis revealed 12 crucial metabolites required to achieve 80% cumulative importance (**Fig. 6D**). These significant metabolites include Alanine, C10-Carnitine, TG(16:0-16:0-20:2), TG(16:1-18:2-18:3), PE(a20:0-20:2), C10:1-Carnitine, Octacosanoic acid(28:0), C14:1OH-Carnitine, C8:1-Carnitine, Asparagine, C0-Carnitine, and d18:0 CE.

Following that, OPLS-DA analysis was carried out using the aforementioned 12 metabolites (**Fig. 6E**). The results, as shown in **Fig. 7**, revealed an AUC of 0.963 (P value = 0.03) for distinguishing CMP vs Other, an AUC of 0.98 (P value = 0.03) for distinguishing IAS vs Other, and an AUC of 0.84 (P value = 0.05) for distinguishing TOF vs Other. These findings indicate promising discriminatory capabilities of the selected metabolites in identifying different CHD subtypes when compared to the other groups.

**Figure 7.** CHD Subtyping modeling with targeted metabolomic profiling analysis of newborn DBS samples. A) Confusion matrix. B) AUC curves to demonstrate the performance to diagnose CHD subtypes.

## Discussion

### Summary of main findings

To explore the identification of CHD metabolic biomarkers using dried blood DBS samples, we began by verifying the reliability of the metabolic profiling from these samples, even after years of storage. This was achieved through correlation analysis of the absolute quantitated metabolite concentrations from this study and the CDPH database records. Subsequently, utilizing global and targeted LC-MS/MS metabolomic profiling methods, we further analyzed the significant metabolic pathways associated with CHD and its subtypes.

In the process, we established models for CHD diagnosis and subtyping to validate the effectiveness of the CHD-associated metabolic patterns. Our findings revealed distinct metabolic differences in significant pathways and unique metabolic patterns related to both CHD and its subtypes. Furthermore, using targeted metabolomics, we identified 3 biomarker metabolites for CHD diagnosis and 12 biomarker metabolites for CHD subtyping. These results support the hypothesis that serial LC-MS profiling of DBS metabolites from newborns may hold potential in accurately assessing CHD and offer a non-invasive and cost-effective method for CHD detection.

Overall, this study sheds light on the potential of utilizing metabolic profiling from DBS samples as a precise and efficient means of CHD diagnosis, paving the way for improved early detection and management of this condition.

### The stability of amino acid and acyl carnitines in DBS

In this study, we utilized CHD DBS samples obtained from the CDPH Biobank, an invaluable resource for research due to its extensive collection of biological specimens suitable for various types of investigations, including epidemiological, genetic, and biomarker research. However, concerns regarding the stability and quality of samples stored in biobanks over extended periods have been raised, limiting retrospective analyses using stored DBS.

To address this concern and ensure the accuracy and reliability of metabolomic studies, we focused on assessing the stability of acylcarnitines and amino acids in DBS stored in freezers after newborn screening. Our research aimed to shed light on the long-term stability of these metabolites in stored specimens.

The encouraging findings of our study revealed excellent stability for both amino acids and acyl carnitines, even after up to 15 years of storage. This discovery provides reassurance that dried blood specimens stored in the CDPH biobank freezers can be reliably used for metabolomic studies, even following prolonged storage. This insight enhances the confidence in utilizing the biobank’s stored samples for future investigations, contributing to the advancement of CHD and other related research areas.

### CHD biomarker biological implications

Informative alterations in plasma metabolite concentrations related to specific metabolic pathways have been found to be associated with the CHD pathogenesis and pathophysiology^27^. A notable example is the significant involvement of fatty acid metabolism in hypertrophied newborn hearts, where it plays a crucial role in energy generation^28^. In our investigation, we discovered a significant association between the arachidonic acid (AA) metabolism pathway and the pathophysiology of various CHD subtypes, including TOF, IAS, and CMP (**Fig. 5A**). The AA metabolism pathway is intricate, involving the metabolism of arachidonic acid, an omega-6 fatty acid, by enzymes like cyclooxygenases (COX), lipoxygenases (LOX), and cytochrome P450 (CYP) enzymes. This pathway gives rise to diverse bioactive lipid mediators, such as prostaglandins, thromboxanes, leukotrienes, and epoxyeicosatrienoic acids (EETs), which play essential roles in various physiological and pathological processes^29^.

Previous studies have demonstrated that dysregulation of the AA metabolism pathway is linked to the development and progression of CHD. For instance, in TOF, a common CHD type, there is evidence of increased oxidative stress and inflammation^30, 31^, leading to potential dysregulation of the AA metabolism pathway. Such dysregulation could result in the overproduction of vasoconstrictive and pro-inflammatory mediators, such as thromboxane A2 and leukotriene B4, contributing to pulmonary hypertension and exacerbating right ventricular dysfunction^32, 33^. Similarly, in inherited arrhythmia syndromes like long QT syndrome and Brugada syndrome, genetic mutations in ion channel genes can alter AA metabolism and lead to the production of EETs. These EETs can modulate ion channel activity, affecting cardiac repolarization and leading to arrhythmias and sudden cardiac death^34–36^. In cardiomyopathies such as hypertrophic cardiomyopathy and dilated cardiomyopathy, elevated inflammation and fibrosis have been observed, potentially contributing to the dysregulation of the AA metabolism pathway^37, 38^. Consequently, the overproduction of pro-inflammatory and pro-fibrotic mediators, such as prostaglandin E2 and leukotriene B4, may play a role in cardiac remodeling and dysfunction. Overall, our findings highlight the relevance of the AA metabolism pathway in CHD pathogenesis and shed light on its potential as a target for further research and therapeutic interventions.

Pathway enrichment analysis conducted on the global metabolic profiling revealed a particularly significant change in the linoleic acid (LA) metabolic pathway in TOF and IAS, while no substantial change was observed in CMP (**Fig. 5A**). It’s worth noting that the LA pathway and AA pathway are closely interrelated. The metabolism of linoleic acid involves a series of enzymatic reactions that ultimately lead to the production of arachidonic acid (AA) and its derivatives, including prostaglandins and leukotrienes, which play pivotal roles in inflammation, cardiovascular function, and other physiological processes. Recent studies have indicated that the linoleic acid pathway may play a role in the development of coronary heart disease [39]. Specifically, research suggests that mutations in the gene responsible for coding delta-6 desaturase (D6D), the enzyme responsible for converting linoleic acid to arachidonic acid, could be associated with an increased risk of coronary heart disease^39^. Furthermore, LA metabolism may also interact with other molecular pathways involved in cardiac development, such as the Wnt signaling pathway, which has been implicated in CHD pathogenesis^40–42^. Although the exact mechanisms by which dysregulation of the LA metabolism pathway contributes to CHD are not yet fully understood, the linoleic acid metabolism pathway has emerged as a potential contributor to the pathogenesis of congenital heart disease. These findings highlight the importance of further research into this metabolic pathway and its potential implications in CHD development and progression.

Monoradylglycerols (MG) are a type of lipid molecule characterized by a glycerol backbone with a single fatty acid molecule attached to it. These lipids are commonly found in various foods, particularly in processed foods that contain added fats and oils. Currently, there is no direct evidence to suggest that MG are directly linked to the development of CHD. However, studies have demonstrated that a diet high in saturated fats, often found in processed foods containing MG, may indeed increase the risk of developing CHD. High-fat diets in expectant mothers have been shown to enhance the expression of placental mRNA associated with the arachidonic acid (AA) metabolism pathway^43^. This can lead to elevated fetal arachidonic acid levels due to maternal arachidonic acid transfer, resulting in an upregulation of fetal VEGF expression^44^. Consequently, an overexpression of VEGF in the myocardium may impede the process of epithelial-mesenchymal transformation, which is crucial for endocardial cushion formation. Such impairment of endocardial cushion formation can trigger defects in septation and outflow tract development^45^. In this context, Smedts et al. reported that a high maternal intake of saturated fat is associated with a higher likelihood of developing outflow tract defects^46^. Furthermore, these researchers found that infants born to mothers with abnormal lipid profiles have twice the risk of CHD^47^. While MG themselves may not be directly linked to CHD, their presence in processed foods that are high in saturated fats may contribute to an increased risk of developing CHD over time. In conclusion, while MG are not directly implicated in CHD development, their association with processed foods high in saturated fats underscores the importance of dietary considerations in reducing the risk of CHD. Adopting a balanced and healthy diet may help mitigate potential risks associated with the consumption of saturated fats and processed foods.

The QHQ pathway is a vital metabolic pathway responsible for maintaining cellular homeostasis and safeguarding cells against oxidative stress. Its primary function involves regulating the redox balance in cells by converting quinones to hydroquinones, which helps control oxidative stress. Oxidative stress arises when there is an imbalance between reactive oxygen species (ROS) production and the cellular antioxidant defense system. Excessive ROS production can lead to oxidative damage of cellular components, such as DNA, proteins, and lipids, ultimately resulting in cellular dysfunction and cell death. This oxidative stress condition has been implicated in the pathogenesis of various diseases, including CHD. Prior studies have highlighted the involvement of oxidative stress in the development of heart malformations, impaired cardiac function, and other complications associated with CHD^48^. Both cyanotic and acyanotic congenital heart diseases have been shown to induce a pro-oxidant state in the human body, leading to increased levels of total antioxidants, oxidants, and the oxidative stress index in affected children^49, 50^. In our study, we observed that the QHQ pathway showed significant differences only in CMP patients (**Fig. 5B**). However, further research is warranted to fully comprehend the underlying mechanisms by which oxidative stress contributes to CHD development. Additionally, identifying potential markers for the early diagnosis of this condition could be crucial in advancing CHD management and treatment strategies. In summary, the QHQ pathway’s involvement in cellular redox regulation and oxidative stress highlights its potential significance in CHD pathogenesis. Understanding its role could provide valuable insights into the development and progression of CHD and pave the way for novel therapeutic interventions and diagnostic approaches.

Furthermore, our study involved targeted metabolomics to profile amino acids, acylcarnitines, and lipids, given their central role in the pathogenesis of heart disease. Carnitine and acylcarnitines play critical roles in energy metabolism, especially in the transport of long-chain fatty acids into the mitochondria for energy production. Infants with CHD are particularly susceptible to energy imbalances, and although most of them have normal weight at birth, they often experience nutritional and growth problems in early infancy^51^. This has spurred growing interest in investigating the potential of carnitine and acylcarnitine for the diagnosis and management of CHD. Several studies have reported decreased levels of carnitine and acylcarnitines in patients with CHDs, particularly those with ventricular overload or pulmonary hypertension^52, 53^. These decreases may indicate impaired cardiac metabolism and energy production, which can contribute to the development of cardiomyopathy and other complications. Beyond carnitine and acylcarnitines, Bahado-Singh et al. conducted a study examining phosphatidylcholine (PC) levels in CHD patients. PC is a major component of cell membranes and is vital for cell structure and function. In the context of CHD, disturbances in tissue remodeling may impact the rate of cell membrane synthesis and destruction, leading to abnormal choline and PC metabolism. Their study revealed significant abnormalities in phosphatidylcholine (PC) and lipid levels in the serum of pregnant women carrying CHD fetuses during the first trimester. Additionally, disturbances in carnitine levels and lipid synthesis were identified, potentially linked to single-carbon metabolism through choline^54^. Consistent with these studies, our research unveiled statistically significant alterations in acylcarnitine and PC levels in DBS samples of CHD patients compared to those of healthy children. Specifically, we identified C12-carnitine, C14:1 carnitine, and PC(d16:1-22:3) as potential diagnostic markers for CHD. These findings provide valuable insights into the metabolic changes associated with CHD and may offer promising avenues for improved diagnosis and management of this condition.

#### Limitations

While our study shows promising potential for metabolomics in CHD newborn screening, it is essential to acknowledge several limitations that may influence the interpretation of our findings. One of the primary limitations is the small sample size used in our study, which may limit the representativeness of our results to a broader population. To enhance the reliability and generalizability of our findings, future research should strive to include larger cohorts, allowing for more robust statistical analysis and validation of the identified biomarkers. Furthermore, it is worth noting that our study was conducted at a specific location, California, USA, which could introduce potential biases related to sample collection, processing, and data analysis. To overcome this limitation and increase the external validity of our findings, conducting multicenter studies in different regions with diverse populations would be highly beneficial. Another important limitation of our study is the lack of consideration for potential confounding factors that may influence the identified biomarkers. Factors such as prenatal exposures, maternal health, and perinatal variables could have an impact on the metabolic signature of CHD. Future research should take these confounders into account to gain a more comprehensive understanding of their influence on metabolomic profiles in CHD. In conclusion, while our study provides valuable insights into the potential of metabolomics in CHD screening, it is crucial to recognize and address these limitations in future investigations. By doing so, we can further enhance the reliability and applicability of metabolomics in early CHD detection and potentially improve patient outcomes.

## Conclusion

Newborn screening using tandem mass spectrometry has been widely adopted as a public health strategy in developed countries to detect inborn errors of metabolism. However, its sensitivity for detecting critical congenital heart disease (CHD) remains relatively low, necessitating the urgent exploration of CHD-related biomarkers. In our study, we have made significant progress in this area. Our research has demonstrated the excellent stability of amino acids and acyl carnitines in dried blood spots (DBS) even after extended storage of up to 15 years. Leveraging this stability, we identified dysregulated metabolic pathways in CHD patients, including alterations in lipid metabolism, energy metabolism, and oxidative stress pathways. Furthermore, through targeted metabolomics, we successfully uncovered potential diagnostic biomarkers, such as acylcarnitines and phosphatidylcholine (PC). These identified biomarker metabolites associated with CHD diagnosis and subtyping hold promising potential as a metabolic signature for CHD newborn screening. This exciting finding opens the door to potential clinical applications of metabolomics in facilitating early CHD detection and assessment, utilizing dry blood spot samples. Moreover, by further investigating the biomarker metabolites and their underlying enriched pathways, we may gain deeper insights into the mechanisms underlying CHD pathophysiology. This can lead to a better understanding of CHD development, which has implications for future research, treatments, and improved patient outcomes. In conclusion, our study highlights the significant progress made in the application of metabolomics for CHD newborn screening and the identification of potential biomarkers. By unraveling the metabolic changes associated with CHD, we can move closer to developing effective screening methods and enhancing our understanding of CHD’s pathogenesis.

## Data Availability

All data produced in the present study are available upon reasonable request to the authors

http://translationalmedicine.stanford.edu

## Acknowledgements

The biospecimens and/or data (Request ID: 1893) used in this study were obtained from the California Biobank Program (CBP) in accordance with Section 6555(a), 17 CCR. The California Department of Public Health is not responsible for the results or conclusions drawn by the authors of this publication.

## Author contributions

Experimental design: S.R. Ceresnac, X.B.Ling

Mass spectrometric assay development: X.B.Ling, Y. Zhang, K.J. Su, J.

Schilling, R.Y. Luo

Sample procurement and data entry: S.R. Ceresnac, X.B.Ling, Y. Zhang

Data analysis: Q. Tang, B. Jin, C.J. Chou, Z. Han, L. Tian,

Clinical Interpretation: S.R. Ceresnac, B.J. Floyd, K.G. Sylvester, H. Chubb, H.J. Cohen, Doff. McElhinney

Manuscript preparation: S.R. Ceresnac, X.B.Ling, Y. Zhang, J.C. Whitin, K.G. Sylvester, H. Chubb, H.J. Cohen, D.B. McElhinney

